# Disproportionality analysis of adverse neurological and psychiatric reactions with the ChAdOx1 (Oxford-AstraZeneca) and BNT162b2 (Pfizer-BioNTech) COVID-19 vaccines in the United Kingdom

**DOI:** 10.1101/2021.09.28.21264245

**Authors:** Santiago Perez-Lloret, Nikolai Petrovsky, Abdallah Alami, James A.G. Crispo, Donald Mattison, Matilde Otero-Losada, Francisco Capani, Daniel Krewski

## Abstract

**Objective:** The information on neurologic or psychiatric adverse reactions to the COVID-19 vaccines is limited. Our objective was to examine the odds of neurological and psychiatric adverse reactions to BNT162b2 (Pfizer-BioNTech) and ChAdOx1 (Oxford-AstraZeneca) COVID-19 vaccines.

**Methods:** We analyzed all Adverse Vaccine Reaction reports to the United Kingdom Medicines and Healthcare products Regulatory Agency between December 9, 2020 and June 30, 2021 that mentioned the BNT162b2 or ChAdOx1 vaccines. We compared the rates of adverse neurological and psychiatric reactions with ChAdOx1 to those with BNT162b2. P-values were obtained by a Bonferroni-adjusted Z-test for proportions.

**Results:** As of June 30, 2021, 53.2 M doses of ChAdOx1 and 46.1 M doses of BNT162b2 had been administered. We extracted information from 300,518 distinct reports. The number of individual adverse neurologic or psychiatric reaction reports were less than 200/M doses administered, except headache which was reported in 1,550 and 395 cases/M doses of ChAdOx1 and BNT162b2, respectively. Compared to BNT162b2, cerebral hemorrhagic or thrombotic events, headaches and migraines, Guillain-Barre syndrome and paresthesias, tremor and freezing, delirium, hallucinations, nervousness, poor sleep quality, and postural dizziness were more frequently reported with ChAdOx1. Reactions more frequently reported with BNT162b2 than ChAdOx1 were Bell’s palsy, facial paralysis, dysgeusia, anxiety, and presyncope or syncope.

**Conclusion:** Significant differences in the neurologic and psychiatric adverse event profiles of the ChAdOx1 and BNT162b2 vaccines may exist, emphasizing the need for additional research. The beneficial and protective effects of the COVID-19 vaccines far outweigh the low potential risk of neurologic and psychiatric reactions.

## INTRODUCTION

The safety and efficacy of COVID-19 vaccines has been studied in short-term clinical trials involving a limited number of participants. Published interim results of Phase III studies have involved: 30,000 participants followed for 28 days following receipt of the mRNA-based mRNA-1273 vaccine (Moderna);[1] 43,000 participants followed for 21 days following receipt of the mRNA-based BNT162b2 vaccine (Pfizer-BioNTech);[2] 22,000 participants followed for 21 days following receipt of the rAd26 and rAd5 vector-based heterologous prime-boost COVID-19 vaccine (Sputnik V, Gamaleya Institute, based on a non-replicating viral vector); and 40,382 participants followed for 14 days following receipt of the WIV04 or HB02 vaccine (China National Biotec Group, a whole inactivated virus vaccine).[3] A pooled analysis of four trials involving the ChAdOx1 (AstraZeneca vaccine, based on a non-replicating viral vector) included 24,422 participants that were followed for at least 14 days.[4] These trials demonstrated that the vaccines were effective in reducing the frequency of symptomatic COVID-19 infection, particularly severe disease, and that adverse events following immunization (AEFIs) were frequent but generally mild to moderate and of reasonably short duration.[5,6] Notwithstanding, clinical trials are not ideal for studying the safety of medical interventions.[7,8] Although they are conducted in tightly controlled environments, their sample size is limited and insufficiently powered to detect anything other than relatively frequent adverse events, and their follow-up periods may be of insufficient duration to detect AEFIs. Real-world pharmacovigilance studies are therefore necessary to identify additional safety issues that may not be observed during vaccine Phase 3 clinical trials.

Vaccines against other infectious agents have been associated with neurologic adverse reactions. For example, infrequent cases of Guillian-Barré Syndrome have been reported following influenza, rabies, tetanus, and polio vaccination. [9,10] Cases of Multiple Sclerosis and Optic Neuritis have also been reported after influenza vaccination.[9] Guillian-Barré Syndrome, Multiple Sclerosis, and Bell’s palsy have been reported after Hepatitis B virus vaccination.[10] Encephalitis, seizures, and sensory disturbances have also been reported after vaccination.[10]

Neurological symptoms (such as headache, myalgia, anosmia, ageusia, impaired consciousness, and psychomotor agitation) have been reported after COVID-19 infection, suggesting viral neurotropism.[11] Therefore, as both mRNA and adenoviral vectors share some properties with the actual virus, particularly their ability to induce expression of the spike protein inside transfected cells rather than the extracellularly as with traditional recombinant protein or inactivated virus vaccines, it is sensible to check for any neurologic and psychiatric AEFIs that may be induced by aberrant tissue expression of spike protein associated with use of such gene therapy vectors.

To address existing knowledge gaps on the safety of COVID-19 vaccines, we examined reports of AEFIs in the United Kingdom linked to use of BNT162b2 and ChAdOx1 COVID-19 vaccines and conducted a disproportionality analysis of neurological and psychiatric AEFIs to evaluate any potential adverse neurological effects of either of these vaccines.

## METHODS

We downloaded information on all suspected adverse reactions that have been reported to the Medicines and Healthcare products Regulatory Agency (MHRA) of the United Kingdom via the Yellow Card scheme for BNT162b2 and ChAdOx1.[12] Reports were gathered between December 9, 2020 and June 30, 2021. We extracted all data on neurologic and psychiatric adverse events from eligible reports (MedDRA 24.0 SOC codes 10029205 or 10037175 respectively).[13] We analyzed individual adverse reactions, as well as the high-level reaction groups to which they belonged. In the MedDRA hierarchy, these correspond to the “Preferred Terms (PT)” for each reaction, which are grouped under the “High Level Terms (HLT)”.

Using the number of adverse reactions reported with BNT162b2 and ChAdOx1 vaccines, we conducted a disproportionality analysis for each reaction and the high-level reaction groups. We excluded adverse reactions with fewer than 5 cases from our analyses. Reporting Odds Ratios (ROR), which in our case represented the ratio of the odds of each event with ChAdOx1 compared to the odds of the event with BNT162b2 (reference group), and their 95% confidence intervals were calculated.[14] Data was analyzed by Z-tests for proportions with a continuity correction. The false positive rate was controlled using a Bonferroni adjustment to compensate for the multiplicity of events analyzed. Critical p-values were 0.0004 and 0.00005 for the HLTs and PTs, respectively. This analysis identified “signals” of adverse reactions potentially related to the use of a specific vaccine.[14] More specifically, 95% confidence interval limits RORs greater than 1 indicated that the AEFI was more frequent with ChAdOx1, whereas RORs 95% confidence interval limits lower than 1 indicated the reaction was more often reported with BNT162b2.

## RESULTS

As of June 30, 2021, 24.6 M and 19.1 M people in the UK received at least one dose of ChAdOx1 or BNT162b2 (Table 1), respectively (total of 43.7 M people vaccinated, accounting for approximately 65% of the UK population).[12] With ChAdOx1, 775,940 AEFIs have been documented in 216,097 reports to MHRA. With BNT162b2, 236,555 AEFIs have been documented in 84,421 reports to MHRA.

**Table 1.** Rates of AEFIs with COVID-19 vaccines

| Reactions | Rate per 1 M<br>ChAdOx1 doses | Rate per 1 M<br>BNT162b2 doses |
| --- | --- | --- |
| Total number of doses administered | 52,300,000 | 46,100,000 |
| Persons that received at least one dose | 24,600,000 | 19,100,000 |
| Persons that received two doses | 21,500,000 | 11,200,000 |
| **High-Level Terms** |  |  |
| *Neurologic* |  |  |
| Acute polyneuropathies | 6.9 | 0.9 |
| Central nervous system hemorrhages and cerebrovascular accidents | 33.7 | 9.4 |
| Cerebrovascular venous and sinus thrombosis | 5.1 | 0.8 |
| Disturbances in consciousness NEC | 190.2 | 80.3 |
| Facial cranial nerve disorders | 17.6 | 13.9 |
| Headaches NEC | 1549.8 | 394.7 |
| Migraine headaches | 144.7 | 41.8 |
| Olfactory nerve disorders | 12.8 | 6.7 |
| Parkinson's disease and parkinsonism | 3.6 | 0.5 |
| Sensory abnormalities NEC | 105.6 | 41.6 |
| Tremor (excl congenital) | 180.5 | 22.2 |
| *Psychiatric* |  |  |
| Deliria | 8.8 | 1.6 |
| Dyssomnias | 12.0 | 2.7 |
| Fluctuating mood symptoms | 1.9 | 1.3 |
| Hallucinations (excl sleep-related) | 19.2 | 3.4 |
| **Preferred Terms** |  |  |
| *Neurologic* |  |  |
| Guillain-Barre syndrome | 6.3 | 0.9 |
| Bell's palsy | 8.9 | 6.5 |
| Anosmia | 5.3 | 3.2 |
| Freezing phenomenon | 2.9 | 0.2 |
| Cluster headache | 13.1 | 3.0 |
| Migraine | 133.7 | 37.7 |
| Facial paralysis | 5.3 | 5.2 |
| Dizziness postural | 39.3 | 10.9 |
| Dysgeusia | 36.8 | 18.8 |
| Tremor | 179.8 | 22.0 |
| Headache | 1486.0 | 377.7 |
| Syncope | 45.4 | 25.0 |
| Paresthesia | 165.0 | 51.6 |
| Presyncope | 10.9 | 7.8 |
| *Psychiatric* |  |  |
| Delirium | 8.7 | 1.6 |
| Hallucination | 17.0 | 2.7 |
| Anxiety | 20.0 | 9.2 |
| Poor quality sleep | 12.0 | 2.7 |
| Nervousness | 10.9 | 2.5 |

We extracted information for 521 reactions, of which 345 (66%) were neurologic and 176 (44%) were psychiatric. These reactions were grouped in 123 distinct HLTs. As shown in Table 1, the most frequently reported AEFI was headache (1550 reports per million doses administered of ChAdOx1 and 395 reports per million doses administered of BNT162b2). The rest of the AEFIs had 200 reports per million doses administered or less (Table 1). We observed significant differences in the proportion of cases reported after ChAdOx1 or BNT162b2 for 19 distinct AEFI PTs and 15 unique HLTs (Table 2).

**Table 2.** “Preferred Terms” and “High-Level Terms” of reactions showing significant differences with the administration of one or the other vaccine.

| Reactions | Unique AEFIs with ChAdOx1 | Unique AEFIs with BNT162b2 | ROR (95% CI) | Z-score |
| --- | --- | --- | --- | --- |
| Total reports | 775,940 | 236,555 | - | - |
| **High-Level Terms** |  |  |  |  |
| *Neurologic* |  |  |  |  |
| Acute polyneuropathies | 361 (0.05%) | 41 (0.02%) | 2.69 (1.94-3.71) | 6.18 |
| Central nervous system hemorrhages and cerebrovascular accidents | 1762 (0.23%) | 433 (0.18%) | 1.24 (1.12-1.38) | 4.01 |
| Cerebrovascular venous and sinus thrombosis | 266 (0.03%) | 37 (0.02%) | 2.19 (1.55-3.09) | 4.52 |
| Disturbances in consciousness NEC | 9947 (1.28%) | 3700 (1.56%) | 0.82 (0.79-0.85) | 10.41 |
| Facial cranial nerve disorders | 919 (0.12%) | 640 (0.27%) | 0.44 (0.40-0.48) | 16.49 |
| Headaches NEC | 81056 (10.45%) | 18197 (7.69%) | 1.40 (1.38-1.42) | 39.43 |
| Migraine headaches | 7567 (0.98%) | 1925 (0.81%) | 1.20 (1.14-1.26) | 7.12 |
| Olfactory nerve disorders | 668 (0.09%) | 309 (0.13%) | 0.66 (0.58-0.75) | 6.07 |
| Parkinson's disease and parkinsonism | 186 (0.02%) | 22 (0.01%) | 2.58 (1.66-4.01) | 4.28 |
| Sensory abnormalities NEC | 5523 (0.71%) | 1920 (0.81%) | 0.88 (0.83-0.92) | 4.96 |
| Tremor (excl congenital) | 9440 (1.22%) | 1024 (0.43%) | 2.83 (2.66-3.02) | 32.98 |
| *Psychiatric* |  |  |  |  |
| Deliria | 458 (0.06%) | 75 (0.03%) | 1.86 (1.46-2.38) | 5.02 |
| Dyssomnias | 630 (0.08%) | 126 (0.05%) | 1.52 (1.26-1.85) | 4.31 |
| Fluctuating mood symptoms | 99 (0.01%) | 58 (0.02%) | 0.52 (0.38-0.72) | 3.93 |
| Hallucinations (excl sleep-related) | 1005 (0.13%) | 159 (0.07%) | 1.93 (1.63-2.28) | 7.79 |
| **Preferred Terms** |  |  |  |  |
| *Neurologic* |  |  |  |  |
| Guillain-Barre syndrome | 332 (0.04%) | 40 (0.02%) | 2.53 (1.82-3.51) | 5.69 |
| Bell's palsy | 467 (0.06%) | 300 (0.13%) | 0.47 (0.41-0.55) | 10.27 |
| Anosmia | 279 (0.04%) | 147 (0.06%) | 0.58 (0.47-0.71) | 5.38 |
| Freezing phenomenon | 153 (0.02%) | 7 (0.01%) | 6.66 (3.12-14.22) | 5.58 |
| Cluster headache | 684 (0.09%) | 136 (0.06%) | 1.53 (1.28-1.84) | 4.55 |
| Migraine | 6993 (0.90%) | 1736 (0.73%) | 1.23 (1.17-1.30) | 7.70 |
| Facial paralysis | 275 (0.04%) | 241 (0.10%) | 0.35 (0.29-0.41) | 12.48 |
| Dizziness postural | 2057 (0.27%) | 504 (0.21%) | 1.24 (1.13-1.37) | 4.39 |
| Dysgeusia | 1924 (0.25%) | 865 (0.37%) | 0.68 (0.62-0.73) | 9.54 |
| Tremor | 9405 (1.21%) | 1012 (0.43%) | 2.86 (2.68-3.05) | 33.08 |
| Headache | 77718 (10.02%) | 17412 (7.36%) | 1.40 (1.38-1.43) | 38.75 |
| Syncope | 2372 (0.31%) | 1152 (0.49%) | 0.63 (0.58-0.67) | 13.09 |
| Paresthesia | 8627 (1.11%) | 2378 (1.01%) | 1.11 (1.06-1.16) | 4.36 |
| Presyncope | 571 (0.07%) | 361 (0.15%) | 0.48 (0.42-0.55) | 11.06 |
| *Psychiatric* |  |  |  |  |
| Delirium | 455 (0.06%) | 75 (0.03%) | 1.85 (1.45-2.36) | 4.96 |
| Hallucination | 888 (0.11%) | 123 (0.05%) | 2.20 (1.82-2.66) | 8.38 |
| Anxiety | 1048 (0.14%) | 423 (0.18%) | 0.75 (0.67-0.85) | 4.86 |
| Poor quality sleep | 626 (0.08%) | 125 (0.05%) | 1.53 (1.26-1.85) | 4.31 |
| Nervousness | 571 (0.07%) | 113 (0.05%) | 1.54 (1.26-1.89) | 4.19 |

Abbreviations: AEFI, Adverse events following immunization; CI, confidence interval; NEC, not elsewhere classified; ROR, reporting odds ratio.

## DISCUSSION

Differences in the frequency of common AEFIs with ChAdOx1 or BNT162b2 vaccines in the UK have been observed.[15] To the best of our knowledge, our study is one of the first attempts to comprehensively analyze the profile of the relatively rare neurologic and psychiatric reactions for these vaccines. We observed significant differences in the frequency of some neurologic and psychiatric reactions in persons exposed to ChAdOx1 or BNT162b2. Cerebral hemorrhagic or thrombotic events, headaches & migraines, Guillain-Barre syndrome (GBS) and paresthesias, tremor and freezing, delirium, hallucinations, nervousness, poor sleep quality, and postural dizziness were all more frequently reported in persons exposed to ChAdOx1. In contrast, Bell’s palsy, facial paralysis, dysgeusia, anxiety, and presyncope or syncope (resulting in altered consciousness) were more frequently reported after BNT162b2 vaccination. The number of neurologic or psychiatric AEFIs reports were in general less than 200 per million doses administered of the vaccines, except for headache which was reported in 1550 and 395 cases per million doses of ChAdOx1 or BNT162b2, respectively.

Analysis of the AEFIs spontaneously reported to the national pharmacovigilance system is a useful approach to detect signals of potential safety issues with drugs or vaccines.[7,8,14] Although such reports are useful to identify potential safety signals for further investigation, they cannot be used by themselves to determine causation. Limitations of our study are that it was not possible to control for imbalances in the characteristics of the vaccinated subjects nor to adjust for other factors potentially associated with the reported adverse reactions. A reporter bias might have been present for some reactions.[16] For example, Bell’s palsy reports may have been spurred by the early findings of an imbalance in reported clinical trials with a higher number of cases of facial paralysis seen in subjects receiving Pfizer or Moderna mRNA vaccines as compared to control subjects.[17]

Our analysis revealed that ChAdOx1 vaccine was associated with more frequent reports of central nervous system thrombotic events as compared to BNT162b2. These findings coincide with those of Cari and colleagues, who compared the rate of events of thrombocytopenia, bleeding, and blood clots after ChAdOx1 or BNT162b2 in the EudraVigilance European database up to April 16, 2021.[18] Adverse reactions caused by thrombocytopenia, bleeding and blood clots were observed in 33 and 151 reports per 1 million doses of BNT162b2 and ChAdOx1, respectively. Cerebral and splanchnic venous thrombosis were also more frequent with ChAdOx1. The physiopathology of cerebral thrombosis after ChAdOx1 is different from that of thrombotic events unrelated to vaccines and is associated with thrombocytopenia.[19] This syndrome, termed “Vaccine-induced Immune Thrombotic Thrombocytopenia”, has been linked to the presence of circulating autoantibodies against platelet factor 4 (PF4).[20] The prothrombotic action of anti-PF4 antibodies may be mediated by the activation of platelets and the engagement of immune effector cells such as monocytes/macrophages and neutrophils.[20] While the physiopathology is not entirely clear, anti-PF4 autoantibody production may be triggered by the inflammation process resulting from the vaccination or could correspond to antibodies triggered by the vaccine cross-reacting with PF4.[21] Our finding of increased frequency of headaches and migraine with ChAdOx1 agrees with the prospective study of Menni and colleagues16 where 282,103 persons immunized with BNT162b2 and 345,280 with ChAdOx1 were followed for 12 days after vaccination in the UK. They observed headaches in 22.8% of ChAdOx1 recipients versus just 7.8% of BNT162b2 recipients, an approximately 3-fold increase.

GBS is a form of polyneuropathy, with acute polyneuropathies and GBS both being reported approximately 2.5 times more frequently with ChAdOx1 than BNT162b2. Cases of GBS have been reported after both ChAdOx1 or BNT162b2.[22–25] In the U.S, 78 cases of GBS have been reported after the use of the adenoviral vector vaccine from Janssen, with a reported frequency of 7.8 cases per million doses,[26] similar to the 6.3 reports per million doses with ChAdOx1, which like the Janssen vaccine is also an adenoviral vector vaccine. Damage to axonal or myelin membranes mediated directly by vaccine virus or vaccine-associated products and cross-reactivity with epitopes on myelin or axonal glycoproteins of peripheral nerves, have been cited as potential mechanisms behind GBS following the administration of vaccines.[27] Further studies are needed to confirm these results and to better understand the potential relationship between acute polyneuropathies including GBS and ChAdOx1.

Some cases of Bell’s palsy have been reported after receipt of either the BNT162b2 or mRNA-1273 vaccine.[22,28,29] Furthermore, the estimated rate ratio of Bell’s palsy among participants of BNT162b2 and mRNA-1273 clinical trials assigned to the vaccine group versus those receiving placebo was 7.0 (p=0.07), suggesting a strong association.[17] Similarly, we observed an increased frequency of Bell’s palsy and some of its symptoms, such as facial paralysis and dysgeusia,[30] after BNT162b2 vaccination. A small, probably underpowered, case-control study failed to find an increased prevalence of Bell’s palsy among participants that had been vaccinated with BNT162b2 compared to unvaccinated controls.[31] However, a population-based study in Hong Kong, that compared the age-adjusted incidence of Bell’s palsy found that vaccination with CoronaVac (an inactivated virus vaccine) or with BNT162b2 was associated with an additional 4.8 or 2.0 cases of Bell’s palsy per 100,000 people vaccinated, respectively.[32] In a nested case-control study using the same database, the authors reported an adjusted odds ratios for Bell’s palsy of 2.385 (95% CI 1.415 to 4.022) for CoronaVac and 1.755 (0.886 to 3.477) for BNT162b2.[32] In the light of the mixed evidence, further research is needed to confirm the potential association between Bell’s palsy and these COVID-19 vaccines.

We also observed an increased frequency of tremor, freezing of gait and other reactions included in the “parkinsonism” category with the ChAdOx1 vaccine. To date, reports of movement disorders cases with COVID-19 and other vaccines are scarce.[33–35] Notwithstanding, videos have circulated on social media about major neurologic adverse events, including movement disorders, after administration of the COVID-19 vaccines, which are thought to represent Functional Neurological Disorders related to psychological distress.[36] The imbalance of cases towards ChAdOx1 observed in the present study, raises some doubts about these outcomes being just functional disorders, as psychological distress would be presumed to be similar among individuals receiving either COVID-19 vaccine. Infection with SARS-CoV-2 and other viruses can cause movement disorders[37,38] and may increase the risk of Parkinson’s Disease.[39] Hence further research on a potential association between Parkinsonian symptoms and ChAdOx1 is warranted.

We also observed an increased frequency of delirium, hallucinations, nervousness, poor sleep quality, and postural dizziness with ChAdOx1, versus anxiety, and presyncope or syncope with BNT162b2. These reactions might reflect psychological distress caused by uncertainty surrounding the COVID-19 pandemic and the risk of adverse reactions with the vaccines. In a recent survey of 2,213 people vaccinated for COVID-19 in Jordan, 33% didn’t believe in long-term vaccine safety and 36% began monitoring their vital signs more often after getting the vaccine.[40] It is not clear why different neurological features were more frequent with BNT162b2 or ChAdOx1 as, again, psychological distress should equally affect people vaccinated with one vaccine or the other.

In summary, while neurologic and psychiatric were relatively infrequent with the vaccines, our disproportionality analysis of reported adverse reactions to ChAdOx1 and BNT162b2 identified some important signals that warrant further examination. Available evidence to date supports the findings of our analysis that ChAdOx1 increases the risk of vaccine-induced immune thrombotic thrombocytopenia syndrome and headaches, while Bell’s Palsy is causally associated with BNT162b2. Further research is needed on any potential associations between GBS and Parkinsonism and tremor with ChAdOx1.

## Data Availability

The data from this study can be accessed at the Medicines and Healthcare products Regulatory Agency website

https://www.gov.uk/government/publications/coronavirus-covid-19-vaccine-adverse-reactions/coronavirus-vaccine-summary-of-yellow-card-reporting

## DECLARATION OF INTERESTS

None

## FUNDING

None

## AUTHOR’S ROLES

Designed the study: SPLL, NP

Accessed the data: SPLL, NP

Analyzed the data: SPLL

Interpreted the results: SPLL, NP, AA, JAGC, DM, MOL, FC, DK

Drafted the manuscript: SPLL

Revised the manuscript: NP, AA, JAGC, DM, MOL, FC, DK

## DATA SHARING

The data from this study can be accessed at the Medicines and Healthcare products Regulatory Agency website: https://www.gov.uk/government/publications/coronavirus-covid-19-vaccine-adverse-reactions/coronavirus-vaccine-summary-of-yellow-card-reporting

## Notes

### Competing Interest Statement

The authors have declared no competing interest.

### Author Declarations

This is a pharmacovigilance dataset, thus approval from an IRB is not needed.

